## Supplementary Notes for "Hotspots of human mutation point to clonal expansions in spermatogonia"

### Supplementary Text

#### 1. Equivalence of *de novo* mutation rate and mutation probability under low mutation rates

In the main text and in the Methods section, the effect of a gene either due to the disease ascertainment in the case of disease-causing genes and variants or due to CES is defined as:

$$\zeta = \frac{P(\mathrm{V}|\mathrm{D})}{P_0(\mathrm{V})} \tag{1}$$

where V is the event of *de novo* mutation either in a gene or in a single position, D is a condition specifying the effect, which can be either ascertainment by disease or V causing clonal expansions in spermatogonia (CES), or both, and $P_0(\mathrm{V})$ is the probability of mutation under the baseline mutation rate.

For a single individual and for any given gene or variant, it holds that $P_0(\mathrm{V}) \ll 1$. And unless $\zeta$ is of the order of $10^3$ for a gene or $10^6$ for a variant or higher (which has not been observed), $P(\mathrm{V}|\mathrm{D}) \ll 1$.

As mutation is a Poisson process with rate given by the mutation rate $\lambda$, it holds that

$$P(\mathrm{G}|\mathrm{D}) = 1 - e^{-\zeta\lambda} \approx \mathbb{E}(\mathrm{V}|\mathrm{D}) = \zeta\lambda, \tag{2}$$

$$P(\mathrm{V}) = 1 - e^{-\lambda} \approx \mathbb{E}(\mathrm{V}) = \lambda \tag{3}$$

i.e. in the regime of low expectations Poisson distribution may be approimated by Bernoulli distribution with the probability equal to the Poisson expectation. If there is a sample of $N$ individuals with mutation rates $\lambda \ll 1$ for V, the expected number of variants under the baseline is $\mathbb{E}(n_{\mathrm{exp}}) = N\lambda$ and the expectation for the number of observed variants is $\mathbb{E}(n_{\mathrm{obs}}) = N\zeta\lambda$. Thus, given (1-3), it holds that

$$\frac{P(\mathrm{V}|\mathrm{D})}{P(\mathrm{V})} \approx \frac{\mathbb{E}(\mathrm{V}|\mathrm{D})}{\mathbb{E}(\mathrm{V})} \approx \frac{N\,\mathbb{E}(\mathrm{V}|\mathrm{D})}{N\,\mathbb{E}(\mathrm{V})} = \frac{\mathbb{E}(n_{\mathrm{obs}})}{\mathbb{E}(n_{\mathrm{exp}})}, \tag{4}$$

and because the ratio $\frac{n_{\mathrm{obs}}}{n_{\mathrm{exp}}}$ is a sample estimate of $\frac{\mathbb{E}(n_{\mathrm{obs}})}{\mathbb{E}(n_{\mathrm{exp}})}$, it holds that

$$\hat{\zeta} = \frac{n_{\mathrm{obs}}}{n_{\mathrm{exp}}} \tag{5}$$

which justifies the usage of the observed-to-expected variant count ratio as a proxy for $\zeta$.

#### 2. CHIP leads to misinterpretation of LOEUF

Genes with significant excess of *de novo* mutations in a disease cohort, but with high LOEUF values, should raise suspicion in studies of their association with the phenotype. Indeed, we constructed the LoF-2 set of putative CES genes this way (see Main Text). However, apart from CES, high LOEUF might be indicative of different biological effects, even if assumptions of strong negative selection against disease-causing genes hold. For example, high LOEUF values may reflect the involvement of these genes in clonal hematopoiesis (CHIP). CHIP increases the frequency of driver mutations in blood[3]; therefore, the analyses of DNA sequenced from blood samples could misinterpret these somatic drivers as germline variants. Due to this effect, LOEUF values may be uninformative about negative selection in the case of CHIP drivers. For example, we identified three CHIP genes with LOEUF $> 0.5$ and significant excess of *de novo* LoF mutations in the ASD/NDD cohort: *DNMT3A*, *PPM1D* and *ASXL1*. Nonetheless, based on the existing medical evidence, we cannot rule out the causal role of these genes in NDD (Suppl. Table S3).

#### 3. Misannotations of PTVs lead to misinterpretations of LOEUF

Both LOEUF and the test on significance of the excess of *de novo* LoF mutations in disease cohorts employed here are based on the assumption that all predicted LoFs have identical functional effects. However, the procedure used to predict LoF is imperfect and introduces functional heterogeneity among LoFs. Most of the time, this misspecification introduces a small amount of noise and just slightly decreases the power of statistical tests. However, in a small number of cases when misclassified LoF have more dramatic functional consequences than LoFs, such misclassification could solely drive association with the disease. In this scenario, *bona fide* LoF could have small phenotypic effect and be under relaxed negative selection, which would manifest as high LOEUF. We encounter two genes in the LoF-2 set (*ODC1*[33] and *PPM1D*[34]), which cause NDD through PTVs bypassed by NMD and therefore act through GoF-like mechanisms. *ODC1* and *PPM1D* have significant excess of predicted LoF in ASD/NDD and LOEUF$> 0.5$. Consistent with the misclassification of GoF PTVs as LoFs, we see that all predicted LoFs in these genes in the ASD/NDD cohort are located in the last or penultimate exons, in line with GoF PTVs described for these genes in the clinical literature[22].

#### 4. Ascertainment of disease-causing variants under arbitrary trait architectures

As mentioned in the main text, mutation rate at a position V may be estimated just from the prevalence of phenotype D it causes. However, a number of assumptions has to be met:
i. Full penetrance of V ($P(\mathrm{D}|\mathrm{V}) = 1$),
ii. D is caused just by V ($P(\mathrm{V}|\mathrm{D}) = 1$),
iii. No transmission or negligible transmission. This assumption ensures that [effectively] all observed variants V are *de novo*.

If a pair $\{\mathrm{V}, \mathrm{D}\}$ satisfies assumptions i-iii, it holds that one estimate of the mutation rate $\hat{\mu}$ is:

$$\begin{aligned} &\hat{\mu} := P(\mathrm{V}) = P(\mathrm{V}) \times 1 = P(\mathrm{V})P(\mathrm{D}|\mathrm{V}) = P(\mathrm{D})P(\mathrm{V}|\mathrm{D}) = P(\mathrm{D}) \times 1 = P(\mathrm{D}), \\ &\therefore \hat{\mu} = P(\mathrm{D}), \end{aligned} \tag{6}$$

where $P(\mathrm{D})$ is by definition the prevalence of D. In the case of incomplete penetrance, (6) becomes a strict inequality and, consequently, $P(\mathrm{D})$ becomes a lower bound for $\hat{\mu}$:

$$\begin{aligned} &P(\mathrm{D}|\mathrm{V}) < 1, \\ &\quad \therefore \hat{\mu} := P(\mathrm{V}) = P(\mathrm{V}) \times 1 > P(\mathrm{V})P(\mathrm{D}|\mathrm{V}) = P(\mathrm{D})P(\mathrm{V}|\mathrm{D}) = P(\mathrm{D}) \times 1 = P(\mathrm{D}), \\ &\quad \therefore \hat{\mu} > P(\mathrm{D}). \end{aligned} \tag{7}$$

Now, if both assumptions (i) and (ii) are dropped, i.e. now $P(\mathrm{V}|\mathrm{D}) \leq 1$ and $P(\mathrm{D}|\mathrm{V}) \leq 1$, we can write $P(\mathrm{V}|\mathrm{D})$ as:

$$P(\mathrm{V}|\mathrm{D}) = \frac{P(\mathrm{V})P(\mathrm{D}|\mathrm{V})}{P(\mathrm{D})},$$
$$P(\mathrm{D}) = P(\mathrm{V})P(\mathrm{D}|\mathrm{V}) + P(\mathrm{V}^c)P(\mathrm{D}|\mathrm{V}^c), \tag{8}$$

where $\mathrm{V}^c$ denotes a complement to the event V occurring with probability $P(\mathrm{V}^c) \equiv 1 - P(\mathrm{V})$. The first term in (8) denotes the part of prevalence of D associated with V, whereas the second term denotes the part of prevalence associated with anything other than V such as other monogenic causes of D, polygenic component, environmental factors etc. Thus, equation (1) in the main text is applicable to arbitrary variants and arbitrary structures of traits given that assumption (iii) is met, and in the case of *de novo* variation studied here (iii) is always satisfied.
